# Early Platelet dynamics during *Staphylococcus aureus* bacteremia is associated with dysregulated cytokine response and predictive of microbial persistence and mortality

**DOI:** 10.1101/2021.08.20.21262372

**Authors:** Rachid Douglas-Louis, Brian V. Lee, Emi Minejima, Mimi Lou, Juliane Bubeck Wardenburg, Annie Wong-Beringer

## Abstract

**Background:** Platelets play key roles in host immune response during sepsis. Microbial persistence and early dysregulated cytokine response predict poor outcomes in patients with *Staphylococcus aureus* bacteremia (SAB). Our objective was to determine the relationship between early platelet trends and cytokine response, microbial persistence and 30-day mortality in patients with SAB.

**Methods:** A two-center observational study was conducted in hospitalized patients with monomicrobial SAB. Electronic medical records were reviewed for pertinent demographic, laboratory, and clinical information. Eligible subjects were grouped by platelet count at onset and Day 4 of SAB: normal platelet (NP, ≥ 150 × 10^9^/L) and thrombocytopenia (TC, < 150 × 10^9^/L). The groups were compared for clinical characteristics and outcomes.

**Results:** 812 patients met inclusion criteria. The median age was 59 years with MRSA accounting for 34% of SAB. The most common comorbidities were hypertension followed by diabetes then renal disease. Thrombocytopenia (TC) occurred in 29% of patients at SAB onset: 15% (n = 120) were mild and 14% (n = 114) were moderate-to-severe (MS). Compared to patients with normal platelet (NP) at SAB onset (n = 578), higher proportion of patients with TC had alcohol use disorder (*p* = 0.015), active malignancy (*p* = 0.002), liver disease (*p* < 0.001), in addition to requiring intensive care unit (ICU) level of care during hospital stay (*p* < 0.001). TC patients had a longer duration of bacteremia (3 days vs 2 days; *p* = 0.008) and higher risk for 30-day mortality (18% vs 6%; *p* < 0.001) overall compared to those with NP. Changes in platelet count from SAB onset to Day 4 differed significantly between those with persistent (PB) versus resolving bacteremia (RB). Notably, patients who had NP at SAB onset but developed new onset of TC by Day 4 had a higher risk for 30-day mortality compared to those who maintained NP at Day 4 (20% vs 4%; *p* < 0.001). Those with recovery of their platelet count from TC to NP by Day 4 (n = 20) had their SAB shortened by one day (2 days vs 3 days; *p* = 0.413) and trended toward lower risk for 30-day mortality (5% vs 22%; *p* = 0.068) compared to those with sustained TC by Day 4 (n = 88).

**Conclusion:** Our results suggest that platelet dynamics during early course of SAB are associated with dysregulated cytokine response and predictive of 30-day mortality. Future studies should be conducted to assess the impact of utilizing platelet count within the first four days of *S. aureus* bacteremia to guide clinical interventions and to further investigate *S. aureus* virulence factors that impair recovery of platelet count in patients with thrombocytopenia.

**Funding:** None

**Research in context:** *Evidence before this study:* Persistent *S. aureus* bacteremia is associated with high mortality and morbidity despite receipt of anti-staphylococcal agents with apparent *in vitro* activity. We have previously reported that early dysregulated host cytokine response biased towards an anti-inflammatory state (high IL-10/TNF ratio) during clinical course of *S. aureus* bacteremia is predictive of persistence and mortality. Additionally, we showed that mortality risk increases by 16% for each continued day with *S. aureus* bacteremia. In recent experimental models, others have shown that platelets play key roles in host immunity and bacterial clearance. Specifically, *S. aureus* virulence factor, α-toxin, induced aberrant platelet aggregation resulting in thrombocytopenia and compromised bacterial clearance from the blood. We performed literature review using PubMed to identify any clinical studies published between January 1, 1996 and April 30, 2021 with the term “thrombocytopenia” AND (“*Staphylococcus aureus*” OR “*S. aureus* bacteremia” OR “persistent bacteremia” OR “duration of *S. aureus* bacteremia” OR “dysregulated immune response”). We identified three related clinical studies published previously: 1) a retrospective study with a main objective of identifying risk factors associated with thrombocytopenia at sepsis onset during *S. aureus* bacteremia, 2) another retrospective study aiming to evaluate the prognostic impact of thrombocytopenia at onset, Day 3, and Day 7 of methicillin-susceptible *S. aureus* bacteremia for predicting mortality, and 3) one prospective study that evaluated the association between ICU admission platelet count and host immune response but did not include infections with *S. aureus*.

*Added value of this study:* To extend the findings from the few earlier clinical studies by performing a more in-depth analysis of the relationship between platelet count, cytokine response, and outcome of bacteremia caused by both methicillin-sensitive and methicillin-resistant *S. aureus* in a large patient cohort. We examined early platelet dynamics by accounting for changes in daily platelet count over the course of initial 7 days of bacteremia which revealed that platelet count during the first three days of *S. aureus* bacteremia was significantly associated with a dysregulated cytokine response and predictive of 30-day mortality. Notably, for every 20 × 10^9^/L drop in platelet count by day 4, the risk of death increased by 25%. Importantly, we observed a significant reduction in the risk of mortality in those with platelet count recovery to normal range by day 4 of bacteremia comparable to those whose platelet count remained normal throughout the course of infection, suggesting a critical window for early therapeutic interventions to mitigate platelet consumption and injury.

*Implications of all available evidence:* Our observations provide strong clinical relevance to the experimental findings implicating the role of *S. aureus* α-toxin in causing platelet injury as well as destruction and sequestration in the microvasculature leading to death in mouse models of bacteremia. The strong link between thrombocytopenia and mortality in *S. aureus* bacteremia support the need for close monitoring of platelets and a more timely and precise approach to *S. aureus* bacteremia management that is based on the host-pathogen interface. Considering the distinct patterns of platelet dynamics observed early during the course of *S. aureus* bacteremia associated with differential mortality risk and the significant improvement in survival among those with platelet recovery by day 4 of bacteremia, follow-up studies should focus on evaluating clinical microbiology procedures that could provide the α-toxin phenotype of the infecting strains with prognostic significance to clinicians and investigating therapeutic agents that could mitigate α-toxin-mediated insults to platelets to improve treatment outcome.

## INTRODUCTION

*Staphylococcus aureus* is a leading cause of sepsis and mortality in the United States.^1^ We have previously shown that, despite receipt of at least three days of anti-staphylococcal therapy with apparent *in vitro* activity, persistent *S. aureus* bacteremia (SAB) occurs in over a third of patients.^2^ Additionally, mortality risk increases by 16% for each continued day with SAB.^3^ Early dysregulated host immune response during clinical course of SAB is predictive of persistence and mortality.^2^ Furthermore, patients with a more pronounced immune response, defined by cytokine levels falling within the upper quartile groups, were at greater risk of developing persistence and mortality.^4^ In addition to thrombosis and hemostasis, recent evidence points to platelets playing key roles in inflammation and infection.^5^ Platelet aggregates in the liver entrap *S. aureus* to facilitate pathogen clearance from the bloodstream.^6,7^ Upon direct interaction with bacteria or their toxins, platelets become activated and secrete microbicidal proteins which can directly kill bacteria, mediate recruitment of other immune cells by chemotaxis, and enhance macrophage activities.^7,8^ Specifically, *S. aureus* secretes α-toxin, a major virulence factor, that mediates pathogenic functions in sepsis by interacting with the widely expressed host cell receptor, a disintegrin and metalloproteinase domain-containing protein 10, on platelets and leukocytes. Thus, this process leads to the disruption of endothelial barriers, stimulation of interleukin-1β (IL-1β) secretion by monocytes, macrophages, and neutrophils, and platelet aggregation and consumption, thereby contributing to microvascular and organ dysfunction.^6,8^ Importantly, others have shown in an experimental model of *S. aureus* sepsis that platelet recruitment and aggregation occurred immediately following infection but that α-toxin induced a second harmful phase of aberrant platelet aggregation in the liver (8 hours or later) that resulted in thrombocytopenia and compromised bacterial clearance from the blood.^9^

Thrombocytopenia has been reported as an independent predictor of mortality in critically ill patients^10^ and specifically among subsets of patients with SAB in a few clinical studies.^11,12^ Another study reported a link between thrombocytopenia and elevated plasma levels for IL-6, IL-8, and IL-10 but the causative pathogens were not reported.^13^ However, the trend in platelet counts during the course of SAB and how it impacts outcome is not clear. Here we aim to determine the impact of platelet dynamics and cytokine response early during the course of SAB on outcome. We hypothesize that thrombocytopenia early during the course of SAB predicts persistence and mortality which supports close monitoring and therapeutic targeting of platelets.

## METHODS

### Study population

This study was conducted at two university-affiliated medical centers (625-bed community and 600-bed county) in Los Angeles County, CA, USA. The study protocol was approved by the institutional review boards at each of the study site. Informed consent was waived as this was an observational study. Adult patients with SAB hospitalized between July 2012 and November 2020 were screened. For patients with multiple admissions during the specified study period, only the initial SAB episode was included. Exclusion criteria were: age less than 18 years, receipt of less than 48 hours of active anti-staphylococcal therapy, delayed initiation of active anti-staphylococcal therapy for more than 48 hours following SAB onset, and polymicrobial bacteremia.

### Data collection

Electronic medical records were retrospectively reviewed to obtain pertinent demographic, laboratory, microbiologic, and clinical data and management details: age, sex, comorbid conditions, vital signs, complete blood cell count, comprehensive metabolic panel, presence of sepsis or septic shock, culture and sensitivity, antibiotic treatment, procedure for source control, time to receipt of active anti-staphylococcal therapy, concomitant use of antiplatelet agents, and platelet transfusion. Pitt Bacteremia Score (PBS) was calculated using the worst score within 48 hours before or on the day of the first positive blood culture. Extracted data were recorded in REDCap, a HIPAA-compliant secured electronic database hosted at the University of Southern California (USC).^15^ Trends in daily platelet count during the initial seven days of SAB were evaluated. Remnant blood samples were obtained at SAB onset and day 4 following onset. Samples were centrifuged and stored at -80 degree Celsius (°C) for later analyses. IL-1β, IL-6, IL-8, IL-10, IL-17, tumor necrosis factor (TNF), and interferon-γ (IFNγ) were measured in duplicates using Luminex multiplex assay (Millipore, Billerica, MA) per manufacturer’s instructions.

### Study definitions

Platelet groups were classified as: normal platelet (NP, > 150 × 10^9^/L), thrombocytopenia (TC, < 150 × 10^9^/L). The degree of thrombocytopenia was further categorized as: mild (Mild TC, 100 to 149 × 10^9^/L) or moderate-to-severe (MS TC, < 100 × 10^9^/L).^12^ The sources of bacteremia were grouped relative to mortality risk^16^: low risk (< 10%) sources were intravenous (IV) catheters, urinary tract infection, ear-nose-larynx, gynecologic, and several manipulation-related sources; intermediate risk (10–20%) sources were osteoarticular, soft-tissue, and unknown sources; high risk (> 20%) sources were endovascular, low respiratory tract, intra-abdominal, and central nervous system foci. Persistent bacteremia was defined by positive blood cultures for three days or longer.^3^ Nosocomial SAB was considered as the first episode that occurred at 48 hours or more following hospital admission.^3^ Mortality was assessed for up to 30-days after the first positive blood culture.^4^

### Data analyses

Patients were grouped by their platelet count per the above definitions (NP, TC; Mild TC and MS TC) and compared for clinical characteristics and outcomes. Descriptive analyses were performed using Student t test or Mann-Whitney U test for continuous variables and Chi-square or Fisher’s exact test for categorial variables. A *p*-value < 0·05 was considered statistically significant. A modified regression analysis using error variance was used to identify the incremental risk for death with each incremental 20-unit drop in platelet count using 150 × 10^9^/L as the reference group. To investigate the association between thrombocytopenia and dysregulated host immune response, and account for the variable cytokine measurements, cytokines levels were stratified into quartile groups as described previously.^4^ Univariate and multivariable regression analyses were performed to determine clinical factors predictive of mortality. Statistical analyses were performed using GraphPad Prism version 9·1·2 (GraphPad Software) or SAS version 9·4 (SAS Institute).

## RESULTS

### Patient characteristics associated with thrombocytopenia at onset of SAB

A total of 842 patients admitted between July 2012 and November 2020 were screened. Of those, 812 patients met inclusion criteria. (**Figure 1**). Patients were grouped by their platelet count at SAB onset; 71% had normal platelet count while 15% had mild TC and 14% had MS TC. (**Table 1**) Overall, most (71%) patients were male and the median age was 59 years. The most common comorbidities were hypertension (50%), diabetes (43%), and renal disease (26%). Patients with TC at onset were more likely to present with coronary artery disease (17% vs 9%; *p* = 0·002) and renal disease (31% vs 24%; *p* = 0·043) compared to those with NP. In addition, TC patients were more likely to present with alcohol use disorder (17% vs 12%; OR 1·673, 95% CI 1·114–2·501; *p* = 0·015), active malignancy (15% vs 8%; OR 2·135, 95% CI 1·315–3·385; *p* = 0·002), and liver disease (28% vs 10%; OR 3·591, 95% CI 2·396–5·312; *p* < 0·001), particularly among patients in the MS group (data not shown). MRSA was the causative pathogen in 34% of bacteremia, regardless of platelet count at SAB onset (30% vs 36% in TC and NP groups, respectively; *p* = 0·12). Incidence of nosocomial SAB was higher in the TC group (21% vs 15%; *p* = 0·038) with shorter time from hospital admission to bacteremia onset compared to the NP group (median 5 days, IQR 3–8 vs 8 days, IQR 4–16; *p* = 0·005). Additionally, those with TC at SAB onset were significantly more likely to get infected from a high-risk source (32% vs 17%; OR 2·228, 95% CI 1·564–3·146; *p* < 0·001) and to present with endocarditis as their primary source of infection (17% vs 6%; OR 3·2, 95% CI 1·945–5·158; *p* < 0·001) compared to NP group. Patients with TC at SAB onset were more likely to be critically ill as reflected by their PBS and the presence of septic shock (*p* < 0·001) compared to those with NP (**Table 1**). One third of all patients required ICU level of care during SAB, with higher proportion of patients in the TC group versus those in the NP group (43% vs 29%; *p* < 0·001). Notably, higher proportion of patients in the MS group experienced septic shock compared to those in the mild TC (31% vs 19%; *p* = 0·049) and NP (31% vs 12%; *p* < 0·001) groups respectively (data not shown). The most common choice for initial anti-staphylococcal agents were vancomycin-containing regimens (76%). Linezolid-containing regimens were used in only 5% overall. TC patients were less likely to have source control procedures performed (35% vs 48%; *p* < 0·001) (**Table 1**).

**Figure 1.**
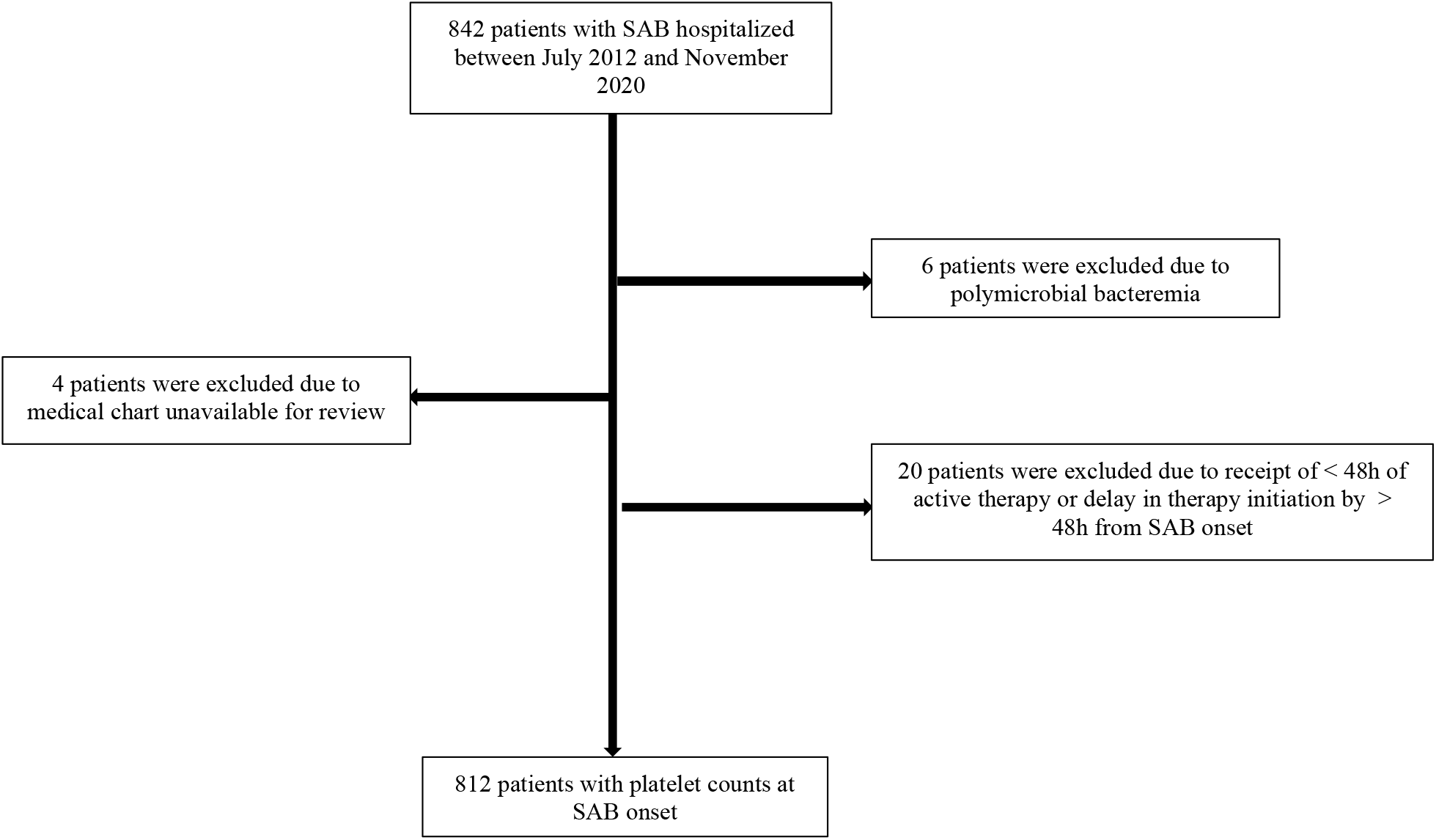
Flowchart of current study. A total of 842 patients were retrospectively screened. Of those, 812 patients met exclusion criteria and were included in the final analysis.

**Table 1.** Demographics, clinical presentation and outcome grouped by platelet count at SAB onset.

| | All patients<br>N = 812 | Normal Platelet<br>($\geq 150 \times 10^9/L$)<br>n = 578 (71) | Thrombocytopenia (<<br>$150 \times 10^9/L$)<br>n = 234 (29) | p-value<sup>a</sup> |
| --- | --- | --- | --- | --- |
| Age, years | 59 (47–70) | 58 (47–69) | 59 (49–72) | 0.164* |
| Male | 578 (71) | 417 (72) | 161 (69) | 0.348 |
| Comorbidities | .. | .. | .. | .. |
| None | 91 (11) | 69 (12) | 22 (9) | 0.225 |
| Active malignancy | 79 (10) | 44 (8) | 35 (15) | 0.002 |
| Alcohol use disorder | 117 (14) | 72 (12) | 45 (17) | 0.015 |
| Coronary artery disease | 94 (12) | 54 (9) | 40 (17) | 0.002 |
| Heart failure | 87 (11) | 58 (10) | 29 (12) | 0.319 |
| Diabetes | 350 (43) | 259 (45) | 91 (39) | 0.137 |
| Hypertension | 403 (50) | 288 (50) | 115 (49) | 0.877 |
| Intravenous drug use | 97 (12) | 73 (13) | 24 (10) | 0.403 |
| Liver disease | 123 (15) | 57 (10) | 66 (28) | < 0.001 |
| Renal disease | 213 (26) | 140 (24) | 73 (31) | 0.043 |
| Dialysis | 143 (18) | 92 (16) | 51 (22) | 0.053 |
| Complicated SAB | .. | .. | .. | .. |
| Endocarditis | 73 (9) | 34 (6) | 40 (17) | < 0.001 |
| Pneumonia | 69 (9) | 46 (8) | 23 (10) | 0.405 |
| Osteomyelitis | 103 (13) | 89 (15) | 14 (6) | < 0.001 |
| Source risk category | .. | .. | .. | .. |
| Low | 166 (20) | 104 (18) | 62 (26) | 0.009 |
| Intermediate | 470 (58) | 373 (65) | 97 (41) | < 0.001 |
| High | 176 (22) | 101 (17) | 75 (32) | < 0.001 |
| Microbiology | .. | .. | .. | .. |
| MRSA | 277 (34) | 207 (36) | 70 (30) | 0.121 |
| Nosocomial | 135 (17) | 86 (15) | 49 (21) | 0.038 |
| Hospital LOS prior to onset,<br>days | 6 (4–13) | 8 (4–16) | 5 (3–8) | 0.005* |
| Severity of Illness | .. | .. | .. | .. |
| Sepsis | 681 (84) | 471 (81) | 210 (90) | 0.003 |
| Severe sepsis | 363 (45) | 209 (36) | 154 (66) | < 0.001 |
| Septic shock | 130 (16) | 72 (12) | 58 (25) | < 0.001 |
| Pitt Bacteremia Score | 1 (0–2) | 1 (0–2) | 1 (0–3) | < 0.001* |
| Source control procedure | 359 (44) | 277 (48) | 82 (35) | < 0.001 |
| Initial active antibiotic regimen | .. | .. | .. | .. |
| Vancomycin-containing regimen | 617 (76) | 443 (77) | 174 (74) | 0.526 |
| Linezolid-containing regimen | 37 (5) | 32 (6) | 5 (2) | 0.040 |
| Concurrent antiplatelets | 200 (25) | 141 (24) | 59 (25) | 0.857 |
| Aspirin | 193 (23) | 135 (23) | 58 (25) | 0.716 |
| Clopidogrel | 35 (4) | 25 (4) | 10 (4) | > 0.999 |
| ICU stay | 269 (33) | 168 (29) | 101 (43) | < 0.001 |
| Duration of SAB, days | 2 (1–4) | 2 (1–4) | 3 (1–5) | 0.008* |
| Persistent SAB | 355 (44) | 237 (41) | 118 (50) | 0.016 |
| 30-day mortality | 75 (9) | 33 (6) | 42 (18) | < 0.001 |

<sup>a</sup>Normal platelet vs thrombocytopenia
Data are n (%) or median (IQR). p-values were determined by Chi-squared or Fisher's exact test or unless indicated otherwise. p-values\* were determined by Kruskal-Wallis test.

### Thrombocytopenia is associated with dysregulated cytokine response

We evaluated the relationship between thrombocytopenia and host cytokine response during SAB in a subset of patients from whom cytokine measurements at SAB onset and Day 4 were available: TNF (n = 127), IL-10 (n = 126), IL-17 (n = 114), IL-6 (n = 94), IL-8 (n = 94), IL-1β (n = 79), IFNγ (n = 75) measurements at SAB onset; and TNF (n = 223), IL-10 (n = 206), IL-8 (n = 192), IL-6 (n = 191), IL-1β (n = 167), IFNγ (n = 158), IL-17 (n = 77) measurements on Day 4. Cytokine measurements were divided into quartile groups as described previously.^4^ Lower platelet count was significantly associated with cytokine response in higher quartiles for both IL-10 and TNF at SAB onset (*p* = 0·001 and *p* = 0·004 respectively) and at Day 4 (*p* < 0·001 and *p* = 0·001 respectively) (**Figure 2A and 2B**).

**Figure 2.**
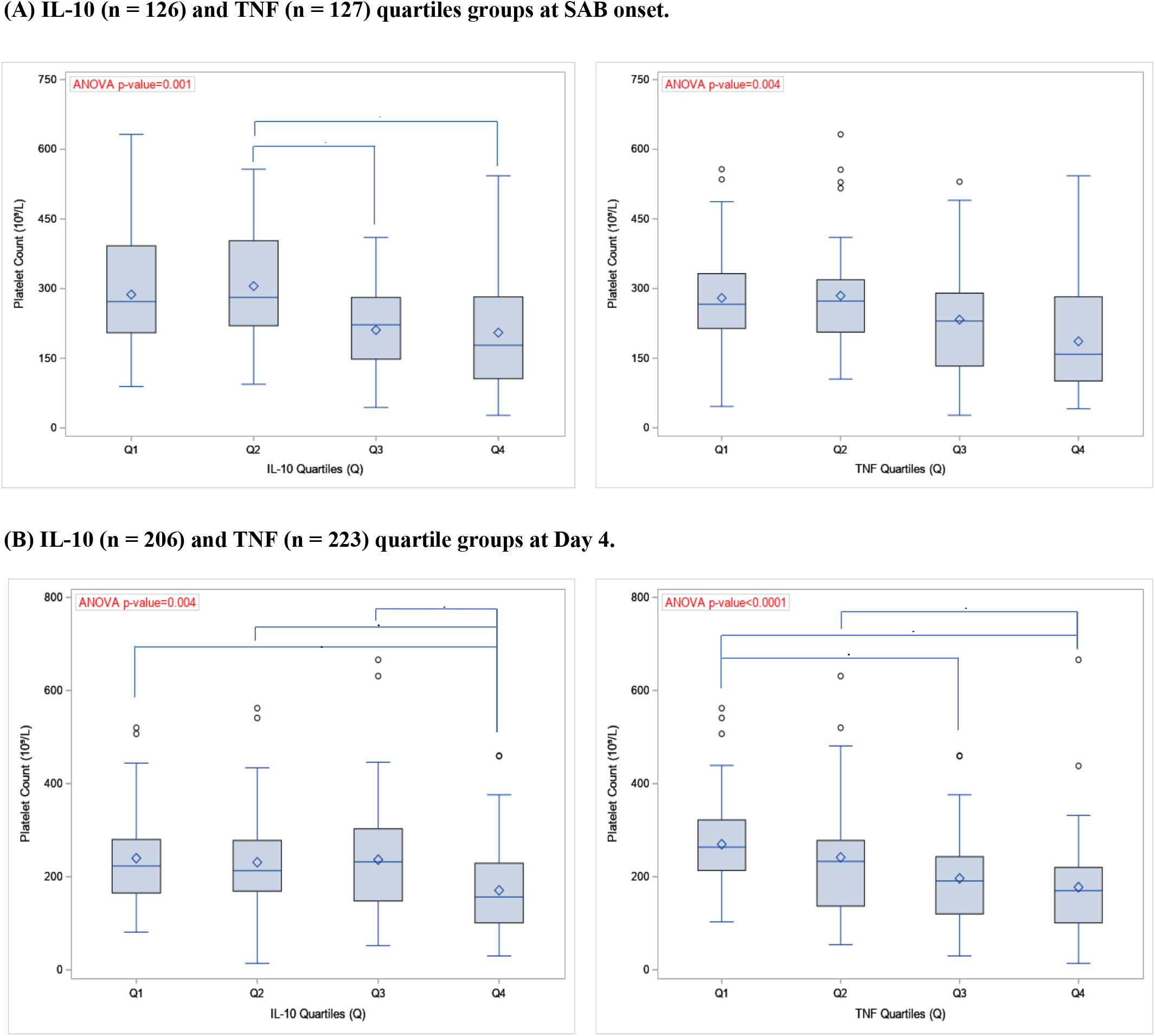
Association of thrombocytopenia with dysregulated cytokine response at onset (A) and Day 4 of SAB (B). Cytokine measurements were divided into quartile groups as described previously.^4^ Lower platelet count was significantly associated with cytokine response in higher quartiles for both IL-10 and TNF at SAB onset (a one-way ANOVA, *p* = 0·001 and *p* = 0·004 respectively) and at Day 4 (a one-way ANOVA, *p* < 0·001 and *p* = 0·001 respectively). *p-value <0.05, **p-value <0.01, adjusted Tukey HSD pairwise comparisons of least squares means. Open circles indicated the extreme observations falling outside the Upper Fence, i.e. the 1.5 (IQR) above 75^th^ percentile.

### Thrombocytopenia increases risk for persistent bacteremia

Bacteremia duration ranged between one and 23 days in our study cohort. The median duration was prolonged by one day for the TC versus NP group (3 days, IQR 1–5 vs 2 days, IQR 1–4; *p* = 0·008) (**Table 1**). After excluding patients with preexisting risks for thrombocytopenia (alcohol use disorder, active malignancy, and liver disease), the risk for persistent bacteremia was significantly greater among those with TC than NP at onset (54% vs 41%; OR 1·978, 95% CI, 1·694–2·528; *p* = 0·012). (**Table 2**)

**Table 2.**
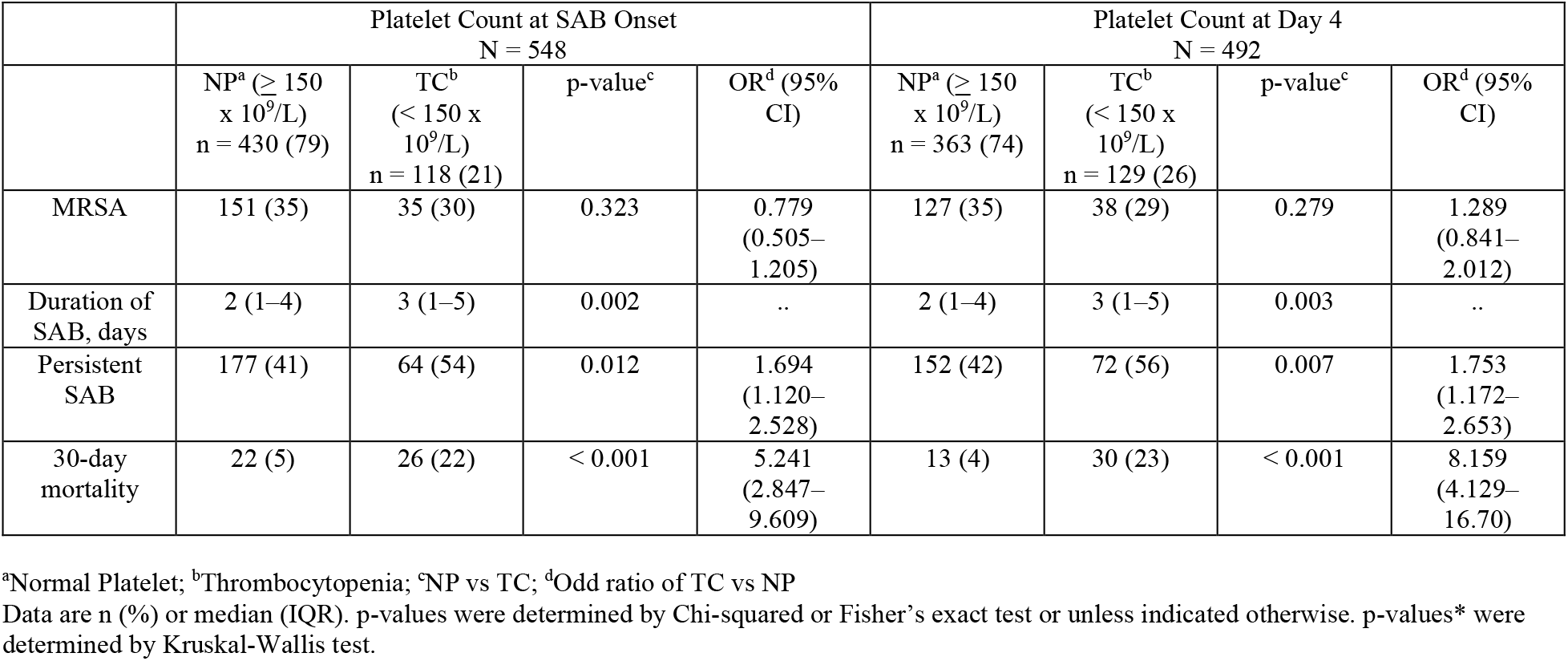
Comparison of infecting strain and clinical outcome grouped by platelet count at SAB onset and on Day 4, excluding those with preexisting contributing factors for thrombocytopenia (e.g., alcohol use disorder, active malignancy, liver disease)

### Thrombocytopenia increases risk of death with greater risk at day 4 than at onset

Overall, 30-day mortality occurred in 9% in this study cohort. (**Table 1**). Compared to NP group, TC at SAB onset was associated with more than three-fold higher risk of death (18% vs 6%; OR 3·613, 95% CI 2·225–5·866; *p* < 0·001). TC compared to NP at day 4 regardless of patients’ platelet count at onset was associated with even greater risk of death (22% vs 5%; OR 5·241, 95% CI 2·847–9·609; *p* < 0·001) (**Table 2**) The association between thrombocytopenia and 30-day mortality was stronger after excluding patients with preexisting risks for thrombocytopenia (alcohol use disorder, active malignancy, and liver disease) than when all patients were considered. (**Table 2**) Based on degree of severity with thrombocytopenia at day 4, mortality was 36% and 12% for the MS TC and Mild TC groups (p=0.002) respectively compared to 4% in the NP group.

### Platelet count dynamics during SAB and impact on mortality

The trend in platelet count over the initial 7-day period following onset of SAB is depicted in **Figures 3A-C**. We observed a biphasic trend in daily platelet count during early phase of SAB where platelet decreased from SAB onset to reach a nadir by Day 2-3 regardless of platelet count at onset. For non-survivors, platelet count decreased early during SAB through Day 4 then continued to remain low over the 7-day period (**Figure 3B**). Notably, we observed a change in platelet count from normal to below normal levels and vice versa during the initial four days of SAB in a subset of 492 patients. Of those, 11% (41/384) of patients with normal platelet count at onset of SAB became thrombocytopenic (NP-TC) by Day 4 whereas 19% (20/108) who were TC at onset had recovery of platelet count to normal level (TC-NP) by Day 4. Of interest, the NP-TC group had more than six-fold greater risk for death (20% vs 3·5%; OR 6·687, 95% CI 2·544–17·14; *p* < 0·001) when compared to those whose platelet count remained normal between SAB onset and Day 4 (NP-NP, n = 343). On the other hand, those in the TC-NP group had reduced lower mortality rate (5% vs 25%; p = 0·068) when compared to those who remained thrombocytopenic (TC-TC) at Day 4. Thus, platelet count on Day 4 appeared to be a determining point of platelet trajectory during the initial 7 days of SAB and is significantly associated with mortality risk. When platelet count on Day 4 was analyzed as a continuous variable, each additional 20 × 10^9^/L drop in platelet count increased mortality risk by 25% compared to those with platelet count ≥ 150 × 10^9^/L (RR 1·25, 95% CI 1·15–1·36; *p* < 0·0001) **(Table 3, Figure 4**).

**Figure 3.**
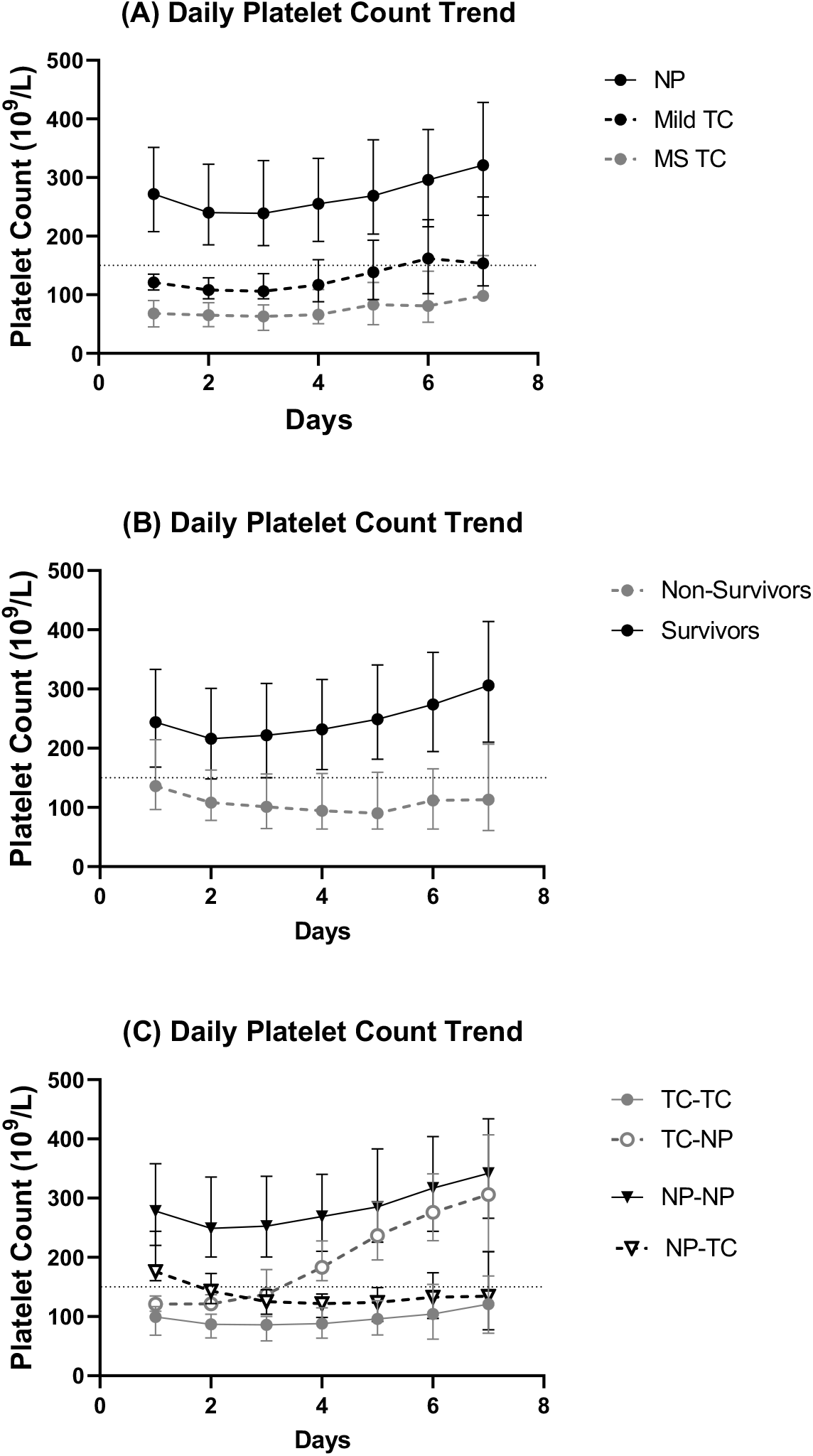
Platelet trends over the first seven days from SAB onset. Groups compared were: (A) Normal Platelet (NP) vs Mild Thrombocytopenia (Mild TC) vs Moderate-to-Severe Thrombocytopenia (MS TC); (B) Survivors vs Non-survivors; (C) Platelet trend for 4 groups based on platelet counts at onset and Day 4: TC-TC (thrombocytopenia at both timepoints), TC-NP (Thrombocytopenia at onset with recovery to normal at day 4), NP-NP (normal platelet count entire course), NP-TC (normal platelet count at onset but thrombocytopenic at Day 4).

**Table 3.** Relative risk for 30-day mortality by platelet count (n = 492). Patients were grouped by platelet count on Day 4 following onset of SAB at 20-unit increment. Patients with platelet count < 50 × 10^9^/L were collapsed into a single group to account for the observed sample size.

| Platelet Count at Day 4<br>(x 10<sup>9</sup>/L) | Total N | Mortality % | Relative Risk (95% CI) | p-value |
| --- | --- | --- | --- | --- |
| > 150 | 363 | 0.3 | Reference | Reference |
| 131–150 | 24 | 4.1 | 2.33 (0.56–9.73) | 0.25 |
| 111–130 | 27 | 3.7 | 1.03 (0.14 - 7.61) | 0.97 |
| 91–110 | 22 | 4.5 | 6.35 (2.49–16.20) | 0.0001 |
| 71–90 | 23 | 4.4 | 9.71 (4.48–21.05) | < 0.0001 |
| 51–70 | 19 | 5.3 | 14.70 (7.42–29.10) | < 0.0001 |
| ≤ 50 | 14 | 7.1 | 7.98 (2.98–21.37) | < 0.0001 |
| Per 20-unit<sup>a</sup> of platelet<br>count | .. | .. | 1.25 (1.15–1.36) | < 0.0001 |

<sup>a</sup>each unit of platelet count = 1 x 10<sup>9</sup>/L

**Figure 4.**
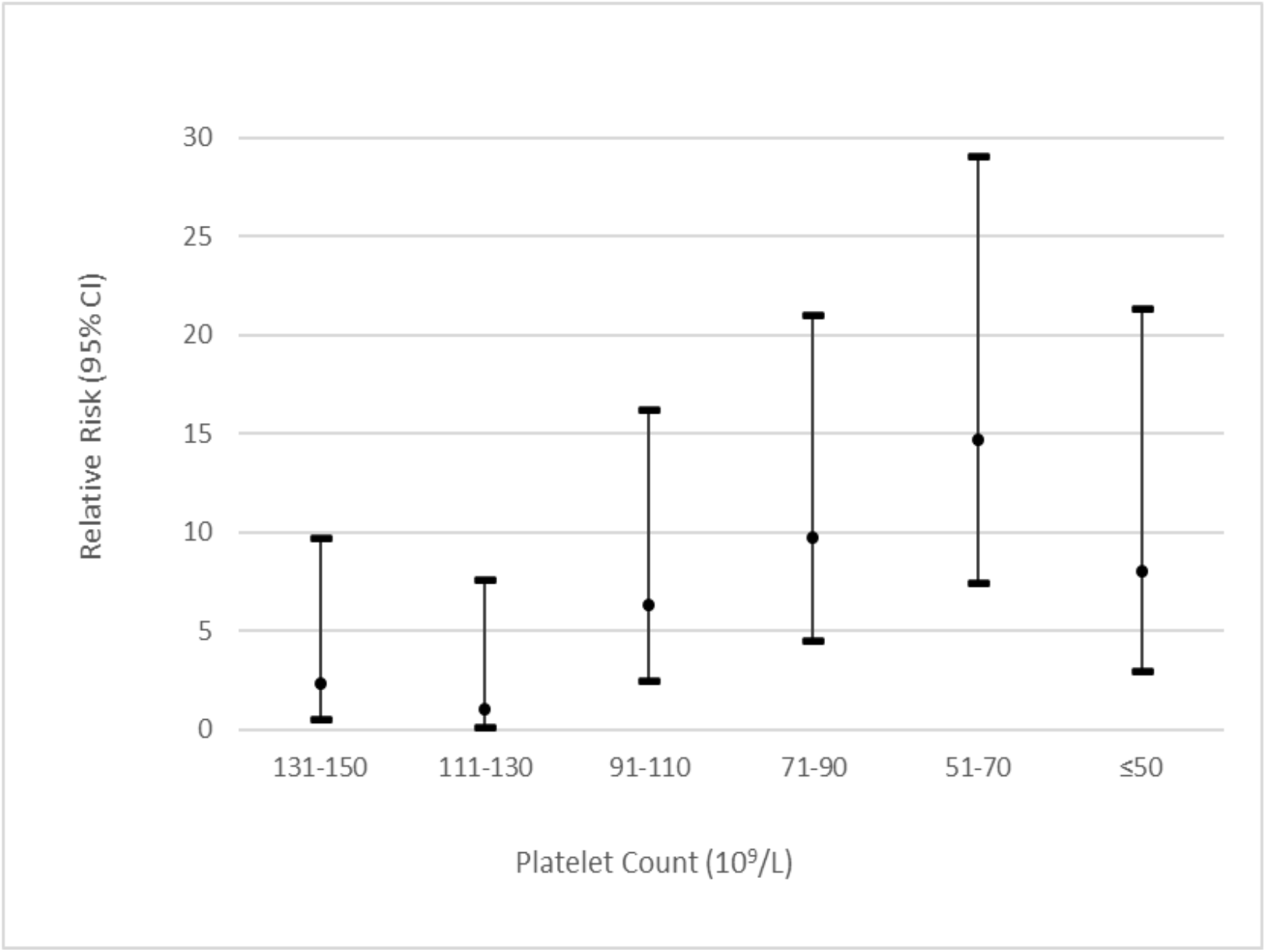
Relative risk (95% confidence interval) of mortality by platelet count (n = 492). Patients were grouped by platelet count on Day 4 following onset of SAB at 20-unit increment. Patients with platelet count < 50 × 10^9^/L were collapsed into a single group to account for the observed sample size.

### Predictors of mortality identified by logistic regression analysis

Univariate and multivariable regression analysis was performed with inclusion of the following variables in the model: age, heart failure, renal disease, source of bacteremia, endocarditis, pneumonia, sepsis, severe sepsis, septic shock, cytokine quartiles, PBS, and source control procedure. By the logistic regression model, the following clinical variables were identified as independent predictors for 30-day mortality: thrombocytopenia at onset (OR 4·078, 95% CI 1·768–9·408; *p* = 0·001), severe sepsis (OR 2·508, 95% CI 1·021–6·152; *p* = 0·04), PBS (OR 1·241, 95% CI 1·086–1·419; *p* < 0·002), and age (OR 1·021, 95% CI 1–1·042; *p* = 0·05) (**Table 4)**. Taking into account the change in platelet count between onset and day 4 of SAB, we performed additional analyses to identify predictors of mortality (**Table 4**). In the sub-model that included only patients who were thrombocytopenic at SAB onset, MRSA infection was significantly associated with 30-day mortality in the group with persistent thrombocytopenia (TC-TC: OR 8·525, 95% CI 2·248–32·335; *p* = 0·002) but not in subgroup whose platelet count recovered to normal level by Day 4 (TC-NP). Among patients who had normal platelets at onset of SAB, independent predictors for mortality were pneumonia (OR 3·106, 95% CI 1·046–9·222; *p* = 0·04), development of thrombocytopenia at Day 4 (OR 2·964, 95% CI 1·001–8·777; *p* = 0·05) and PBS (OR 1·212, 95% CI 1·022–1·438; *p* = 0·03).

**Table 4.** Multivariable predictive models for 30-day mortality for all patients and subgroups based on platelet trends from SAB onset to Day 4.

| Full Model: All patients (N=483; 440 survivors vs 43 non-survivors) |  |  |  |  |
| --- | --- | --- | --- | --- |
| Variables | Odds Ratio Estimates | 95% CI |  | p-value |
| Group TC-TC vs NP-NP | 4.078 | 1.768 | 9.408 | 0.001 |
| Group TC-NP vs NP-NP | 1.768 | 0.269 | 11.636 | 0.55 |
| Group NP-TC vs NP-NP | 2.550 | 0.831 | 7.822 | 0.10 |
| Source risk category, Intermediate vs. High | 0.282 | 0.128 | 0.621 | 0.002 |
| Low vs. High | 0.235 | 0.086 | 0.645 | 0.005 |
| Pitt Bacteremia Score | 1.241 | 1.086 | 1.419 | 0.002 |
| Severe Sepsis (Yes vs. No) | 2.508 | 1.021 | 6.162 | 0.04 |
| Age | 1.021 | 1.000 | 1.042 | 0.05 |

| Submodel: Patients with normal platelet count at onset, N=382; 361 survivors vs 20 non-survivors |  |  |  |  |
| --- | --- | --- | --- | --- |
| Variables | Odds Ratio Estimates | 95% CI |  | p-value |
| NP-TC (vs. NP-NP) – forced in model | 2.964 | 1.001 | 8.777 | 0.05 |
| Pitt Bacteremia Score | 1.212 | 1.022 | 1.438 | 0.03 |
| Age | 1.030 | 0.999 | 1.061 | 0.06 |
| Pneumonia (Yes. vs. No) | 3.106 | 1.046 | 9.222 | 0.04 |

| Submodel: Patients with thrombocytopenia at time of onset, N=108; 85 survivors vs 23 non-survivors |  |  |  |  |
| --- | --- | --- | --- | --- |
| Variables | Odds Ratio Estimates | 95% CI |  | p-value |
| TC-TC (vs. TC-NP) - forced in model | 2.138 | 0.301 | 15.199 | 0.45 |
| Pitt Bacteremia Score | 1.490 | 1.172 | 1.894 | 0.001 |
| Source risk category, Intermediate vs. High | 0.221 | 0.056 | 0.871 | 0.03 |
| Low vs. High | 0.094 | 0.019 | 0.476 | 0.004 |
| MRSA (vs. MSSA) | 8.525 | 2.248 | 32.335 | 0.002 |

NOTE: TC-TC = thrombocytopenia at onset through day 4; NP-NP = normal platelet count at onset through day 4; TC-NP = thrombocytopenia at onset with platelet count recovery to normal limits by day 4; NP-TC = normal platelet count at onset with development of thrombocytopenia by day 4.

## DISCUSSION

Thrombocytopenia during hospitalization is a common hematologic dysfunction in critically ill patients.^17-19^ Past studies have pointed to thrombocytopenia as a marker for sepsis and serious infections that is associated with poor outcomes.^20,21^ In the present study, we found that 29% of patients had thrombocytopenia at onset of SAB and an additional 8% developed thrombocytopenia at Day 4 following onset. We sought to determine the association between thrombocytopenia and dysregulated cytokine response, persistent bacteremia, and 30-day mortality. We observed that patients who developed thrombocytopenia at onset were significantly more likely to be admitted to the ICU and have active malignancy, liver disease, alcohol use disorder, and endocarditis. After exclusion of preexisting contributing factors for thrombocytopenia, a stronger link between thrombocytopenia and the risk for death was observed. Compared to survivors, non-survivors had significantly lower platelet count following SAB onset, which continued to remain low after Day 4. Our findings were consistent with results from Gafter-Gvili and colleagues who reported an association between thrombocytopenia at sepsis onset and 30-day mortality^11^; however, they observed a much higher mortality rate in their study cohort than ours (56% vs 18%). This could be attributed to the difference in our respective study populations in which their study included a higher proportion of patients with severe sepsis, infection with MRSA, and polymicrobial bacteremia. Importantly, we extended the literature by examining daily platelet trend during the initial 7 days of bacteremia and its relationship with persistent bacteremia and 30-day mortality. Forsblom and colleagues have shown that thrombocytopenia at Day 7 following SAB onset was associated with 90-day mortality.^12^ Interestingly, they observed that thrombocytopenia at SAB onset was not associated with 28-day mortality. It is notable that their study included only patients with MSSA bacteremia which may account for the observed differences. It is worth noting that MRSA was a significant predictor for mortality that is linked to the lack of platelet recovery by Day 4 in our multivariable logistic regression model. Our lab has shown that MRSA bloodstream isolates produced higher α-toxin protein levels and hemolytic activity compared to MSSA strains. In addition, α-toxin-mediated hemolytic activity was significantly associated with thrombocytopenia and mortality in patients with bacteremia caused by MRSA but not by MSSA (unpublished data).

In line with published findings of platelet trend in the setting of an acute illness,^22^ we observed a biphasic trend in platelet count during early phase of SAB where platelet decreased from onset of bacteremia reaching a nadir by Day 2-3 then returned to baseline level or higher by Day 7. Notably, a close examination of platelet dynamics during the initial four days of bacteremia indicated that regardless of platelet count at onset, platelet count at Day 4 appeared to be an inflection point indicative of the platelet trajectory with prognostic significance. When platelet count on Day 4 was evaluated as a continuous variable, the risk of death increased by 25% for every 20 × 10^9^/L drop in platelet count. In particular, risk of death in patients with normal platelet counts at SAB onset but became thrombocytopenic by Day 4 approached the risk for death in those who were thrombocytopenic throughout the course of bacteremia (20% vs 25% respectively). On the other hand, despite being thrombocytopenic at onset of bacteremia, risk of mortality was significantly reduced if platelet count recovered to normal range comparable to those whose platelet count remained normal throughout the course of infection (5% vs 3.5% respectively). The latter observation suggests a window of opportunity for therapeutic intervention to protect platelets from destruction or to augment recovery. Of interest, we and others have previously shown that antibiotics have differential effects on α-toxin expression^23^; selected β-lactam agents were shown to induce α-toxin gene expression while some of the protein synthesis inhibitors (e.g., clindamycin, tedizolid, linezolid) were shown to inhibit α-toxin protein production.^24^ Taken together, it is conceivable that precision antibiotic therapy may be prescribed to patients based on measures of pathogen virulence (α-toxin expression of *S. aureus*) and susceptibility by harnessing the dual antimicrobial and anti-virulence potential of antibiotics to improve treatment outcome of *S. aureus* bacteremia.

Recent evidence has suggested that platelets contributed to the dysregulated host immune response during sepsis.^13^ As we have previously reported, a dysregulated cytokine response in TNF and IL-10 levels at onset and on Day 4 of SAB was associated with persistent bacteremia and mortality.^2,13^ Here we found that thrombocytopenia at SAB onset and on Day 4 was significantly associated with an exuberant response in IL-10 and TNF. These findings provide further support for the key role that platelets play in the pathogenesis of sepsis that links to dysregulated cytokine response.

We acknowledge several limitations in our study. Because this was a retrospective observational study, drug-related adverse events, onset of bleeding or other causes of thrombocytopenia could not be fully evaluated. The incidence of disseminated intravascular coagulation, heparin-induced thrombocytopenia may have been underestimated due to the inconsistent documentation in patients’ electronic medical records. The timing of platelet transfusion and post-transfusion platelet count were not captured thus we were uncertain of the impact on daily platelet count. We adjusted our final analyses to limit confounders by excluding patients with preexisting risks for thrombocytopenia; we observed an even stronger link between platelet count and mortality in this patient subset. To our knowledge, this is the first large study that included 812 patients with SAB in whom we have multiple platelet count measures during the course of bacteremia to assess their impact on cytokine response, bacterial persistence and mortality.

Taken together, findings from our study could have an immediate impact on current practice regarding the management of patients with SAB. Platelet count should be closely monitored in patients with SAB particularly during the initial four days of infection and in those infected with MRSA. Concomitant medications with the potential to cause thrombocytopenia should be used with caution depending on their mechanism affecting platelets. Future studies should evaluate the feasibility of measuring *S. aureus* α-toxin phenotype using an agar-based approach to assess α-toxin-mediated hemolysis as part of the routine workflow in clinical microbiology. Clinical impact of administering adjunctive α-toxin-inhibiting or mitigating agents (e.g., clindamycin, ticagrelor, oseltamivir) to protect against platelet injury or depletion on outcome of SAB deserves further investigations.^25^

In conclusion, our findings suggested that thrombocytopenia during early course of SAB was significantly associated with persistent bacteremia and 30-day mortality. Platelet dynamics during the initial four days of infection carried prognostic significance and represented a window of opportunity for therapeutic interventions that deserve to be further investigated.

## Data Availability

Participant data that underlie the results reported in this article after de-identification will be made available upon reasonable request, beginning 9 months and ending 36 months following article publication in a peer-reviewed journal. To gain access, data requestors will need to sign a data access agreement.

## Authors and Contributors

RDL contributed data collection and analysis, wrote the main manuscript text, and prepared tables and figures. BVL contributed data collection and analysis. ML was responsible for statistical analysis. EM supervised study site and contributed to data management. JBW critically revised the manuscript. AWB conceived and designed the study, contributed analysis, critically revised the manuscript. The authors read and approved the final manuscript.

## Acknowledgements

We thank the Clinical Laboratory staff at Huntington Memorial Hospital and Los Angeles County + University of Southern California Medical Center for assistance in collection of bacterial and blood specimens. The content is solely the responsibility of the authors and does not necessarily represent the official views of Huntington Memorial Hospital and LAC + USC Medical Center

